# Library preparation strategy critically impacts RNA virus sensitivity in clinical metagenomics

**DOI:** 10.64898/2026.05.18.26353500

**Authors:** Dominika Stepniak, Bede Constantinides, Mia Weaver, Samantha Treagus, Sam AJ Wilkinson, Samuel Quarton, Mahboobeh Behruznia, Nicola Cumley, John Tyson, Alan McNally, Nicholas J Loman, Steven Pullan, Joshua Quick

## Abstract

Clinical metagenomics uses sequencing for culture-independent identification of pathogens directly from clinical specimens. While a number of protocols claim to be pathogen agnostic, sensitivity for RNA viruses is likely lower than for bacteria or fungi, as it requires additional processing steps including conversion to cDNA. Sequence-independent, single-primer amplification (SISPA) was first described in 1991, yet how it preferentially enriches viral molecules has never been described. Here we propose that single-primer amplification exploits the PCR suppression effect, which selectively amplifies longer viral molecules over shorter host-derived cDNA fragments on the basis of size. This model predicts that any upstream processing step that disrupts fragment length will prevent this enrichment occurring. To test this, we systematically compared two adapter introduction strategies - during cDNA synthesis and via tagmentation - followed by single primer amplification, using the ZeptoMetrix Respiratory Panel 2.1 containing 16 RNA and 3 DNA virus strains. SISPA-based approaches recovered all of the viral genomes in the control, whereas using tagmentation to amplify cDNA recovered none. We then spiked the controls into extracted clinical samples and found that SISPA-based methods performed best in all background settings, however in high-background settings no viral genomes were recovered by any approach. Finally, using a modified SMART-9N protocol, we demonstrated that single-primer PCR is critical to overall performance, indicating that direct tagmentation of cDNA and dual-primer PCR should be avoided in protocols for clinical metagenomics where high sensitivity for RNA viruses is critical. These findings demonstrate that library preparation strategy fundamentally determines RNA virus sensitivity and offer mechanistic insights for protocol optimisation with direct relevance to clinical metagenomics.

## Introduction

Clinical metagenomics for pathogen identification has gained renewed momentum as researchers seek culture-independent methods capable of sequencing pathogens directly from clinical specimens. Potential pathogens include viral, bacterial, fungal, and parasitic organisms with diverse physical properties and genome types. A fundamental challenge in clinical metagenomics is overcoming the high ratio of host to pathogen nucleic acid (Yu et al. 2024; Kim et al. 2024). High background is one of the largest contributors to false negative results in clinical metagenomics (Benoit et al. 2024). Several host depletion strategies have been developed to address this background problem. The inclusion of DNase treatment before extraction (Allander et al. 2001) improves the viral-to-host ratio by degrading free DNA. Filtration and centrifugation (Lewandowska et al. 2015; Liu et al. 2020) have been used to separate cells based on size difference; this is often used in conjunction with DNase treatment because much of the host DNA in clinical samples is extracellular. For bacterial pathogens, saponin-based differential lysis of human cells followed by DNase treatment has proven effective in reducing host by up to 10,000 times (Hasan et al. 2016; Charalampous et al. 2019) although it has been reported to also affect the levels of Gram-negative bacteria (Longhi et al. 2024). Other host depletion methods include osmotic lysis, propidium monoazide (PMA) treatment (Marotz et al. 2018), antibody-based 5mC methylation capture (Feehery et al. 2013) and CRISPR-Cas9 depletion (Gu et al. 2016).

### Protocols for clinical metagenomics

We hypothesise that sensitivity for RNA viruses is determined by the molecular mechanisms underlying library preparation, specifically the conversion of RNA to cDNA followed by adapter introduction and PCR amplification. Two fundamentally different strategies exist for introducing PCR adapters: (1) during cDNA synthesis by incorporating adapter sequences into primers, creating full-length molecules with inverted terminal repeats (ITRs), or (2) by transposase-mediated tagmentation, which fragments cDNA and introduces adapters simultaneously. These approaches interact differently with downstream PCR amplification which for single primer PCR will be subject to the PCR suppression (PS-) effect (Siebert et al. 1995; Dai et al. 2007). The Rapid Metagenomics Protocol (Oxford Nanopore Technologies) has cellular and acellular arms. Saponin/DNase treatment (cellular) and DNase treatment (acellular) are used for host depletion and adapters are introduced during cDNA synthesis (acellular) and via tagmentation (cellular). DNase treatment is performed prior to a total nucleic acid extraction, leaving encapsidated DNA viral genomes intact.

Through a mechanism of random priming during the PCR, it allows the sequencing of DNA and RNA viruses simultaneously (Wright et al. 2015). Pan-microbial (Alcolea-Medina et al. 2024) is a single arm protocol which uses differential lysis/DNase treatment for host depletion and tagmentation for adapter introduction. UCSF CSF mNGS assay (Miller et al. 2019) has DNA and RNA arms which use methylation capture (DNA) and DNase treatment (RNA) host depletion and uses tagmentation for adapter introduction for both. The untargeted metagenomics protocol for CSF and tissues from sterile sites (Atkinson et al. 2023) has DNA and RNA arms which use methylation capture (DNA) and ribosomal RNA depletion for tissue samples and ligation-based library preparation.

### The PCR suppression effect

Single-primer PCR amplification, where all products contain the same adapter sequence at both ends, is subject to the PS-effect. After denaturation single-stranded DNA molecules with inverted terminal repeats can form intramolecular ‘panhandle’ or hairpin structures rather than annealing to primers. This suppresses exponential amplification of molecules that form stable panhandles. Critically, suppression efficiency inversely correlates with fragment length: shorter fragments form panhandles more readily and are strongly suppressed, while longer fragments remain accessible to primer binding for longer and so amplify more efficiently. The effect has been shown to be tunable with adapter length, GC content and primer concentration, affecting the average length of complex PCR products (Shagin et al. 1999). This length-dependent enrichment can be exploited for viral enrichment. Total RNA is dominated by ribosomal RNA (∼85%) (Karpinets et al. 2006) and transfer RNA (10–15%). While large ribosomal RNA subunits can reach kilobase lengths, the most abundant RNA species by copy number—transfer RNA and small ribosomal RNAs— are considerably shorter (76–160 nucleotides) than intact viral genomic RNA molecules (typically 7–30 kilobases for respiratory viruses). Sample handling and nuclease activity result in partial fragmentation of all RNA species, but this length differential enables PCR suppression-mediated enrichment of viral targets during single-primer PCR amplification.

### Random-primed cDNA synthesis approaches for viral metagenomics

This principle underlies sequence-independent single-primer amplification (SISPA) methods (Reyes & Kim 1991; Froussard 1992) which include the Round A/B approach (Wang et al. 2003) which uses tagged random primers to incorporate ITRs during cDNA synthesis. Round A generates random-primed, double-stranded cDNA through primer extension using reverse transcriptase and Sequenase—a modified T7 DNA polymerase with reduced 3′ to 5′ exonuclease activity (Bohlander et al. 1992). Round B performs single-primer PCR in which longer viral molecules are preferentially amplified due to reduced suppression. SMART-9N employs the SMART (Switching Mechanism at the 5′ end of RNA Template) (Zhu et al. 2001) system to generate random-primed first-strand cDNA in a single-tube reaction, followed by single-primer PCR amplification (Claro et al. 2021). While slower than Round A/B due to rate-limiting template-switching activity, SMART-9N offers a practical advantage by combining both 3′ and 5′ adapter incorporation into a single reaction, which minimises hands-on time and maintains the cDNA in a smaller reaction volume. This volume reduction increases the effective concentration of viral genome copies entering the subsequent PCR step, enhancing sensitivity particularly for low-titer clinical samples.

### Tagmentation-based library preparation

Tagmentation-based approaches have become the first choice for NGS library preparation. Exploiting the cut-and-paste activity of transposase enzymes, they combine fragmentation and adapter ligation into a single reaction reducing time and cost. The transposome in the PCR Barcoding Kit (Oxford Nanopore) contains a single sequence allowing the strand-transfer complex to be amplified with a single PCR primer. The Nextera family of kits (Illumina) contain a mixture of A- and B-adapters, meaning 50% of strand-transfer complexes can be amplified using two barcoded PCR primers. Transposase-based kits using single-primer PCR will be subject to PS-effect based enrichment whereas others will not. Transposase-based methods are sensitive to input DNA quantity. Insert size distribution depends on the mass ratio of input DNA to transposase complex: low input produces a smaller distance between insertion sites, while high input produces a larger distance between insertion sites. The exception to this is Nextera Flex which is bead-based meaning the spacing of the transpososomes on the surface of the bead determines the insert size rather than DNA quantity. For clinical metagenomics, where effective host depletion reduces input material and where some sample types have very low biomass, this presents a challenge.

We investigated whether adapter introduction strategy—during cDNA synthesis versus post-cDNA synthesis tagmentation— in conjunction with different amplification strategies impacts RNA virus sensitivity in metagenomic sequencing. Using the Zeptometrix Respiratory 2.1 panel as a reference standard, we compared SISPA-based approaches (SMART-9N and Round A/B) with a tagmentation-based approach, followed by single primer amplification. We used the same Zeptometrix panel spiked into host-depleted clinical sample matrices of high, medium and low backgrounds to determine input sensitivity. Finally, we compared the performance of single and dual primer PCR using a modified SMART-9N approach.

## Methods

### Overview of experiments

#### Impact of adapter introduction strategy

For this investigation we used two cDNA synthesis methods used in SISPA-based protocols (Round A and SMART-9N) both of which introduce adapters as inverted terminal repeats (ITRs). SMART-9N generates a 5’ full-length, DNA/RNA heteroduplex via the template-switching mechanism and Round A generates double-stranded cDNA using a combination of reverse transcriptase and DNA polymerase. These are referred to as ‘SMART-9N/Full’ and ‘Round A/Full’. To assess transposase-mediated adapter introduction ‘SMART-9N/Full’ and ‘Round A/Full’ were tagmented (see ‘Tagmentation’ subheading below) to create ‘SMART-9N/Tag’ and ‘Round A/Tag’. These were amplified by single-primer PCR using ‘NEB’ or ‘RLB’ or both ‘NEB&RLB’ producing a total of eight conditions **(Figure 1)**.

**Figure 1.**
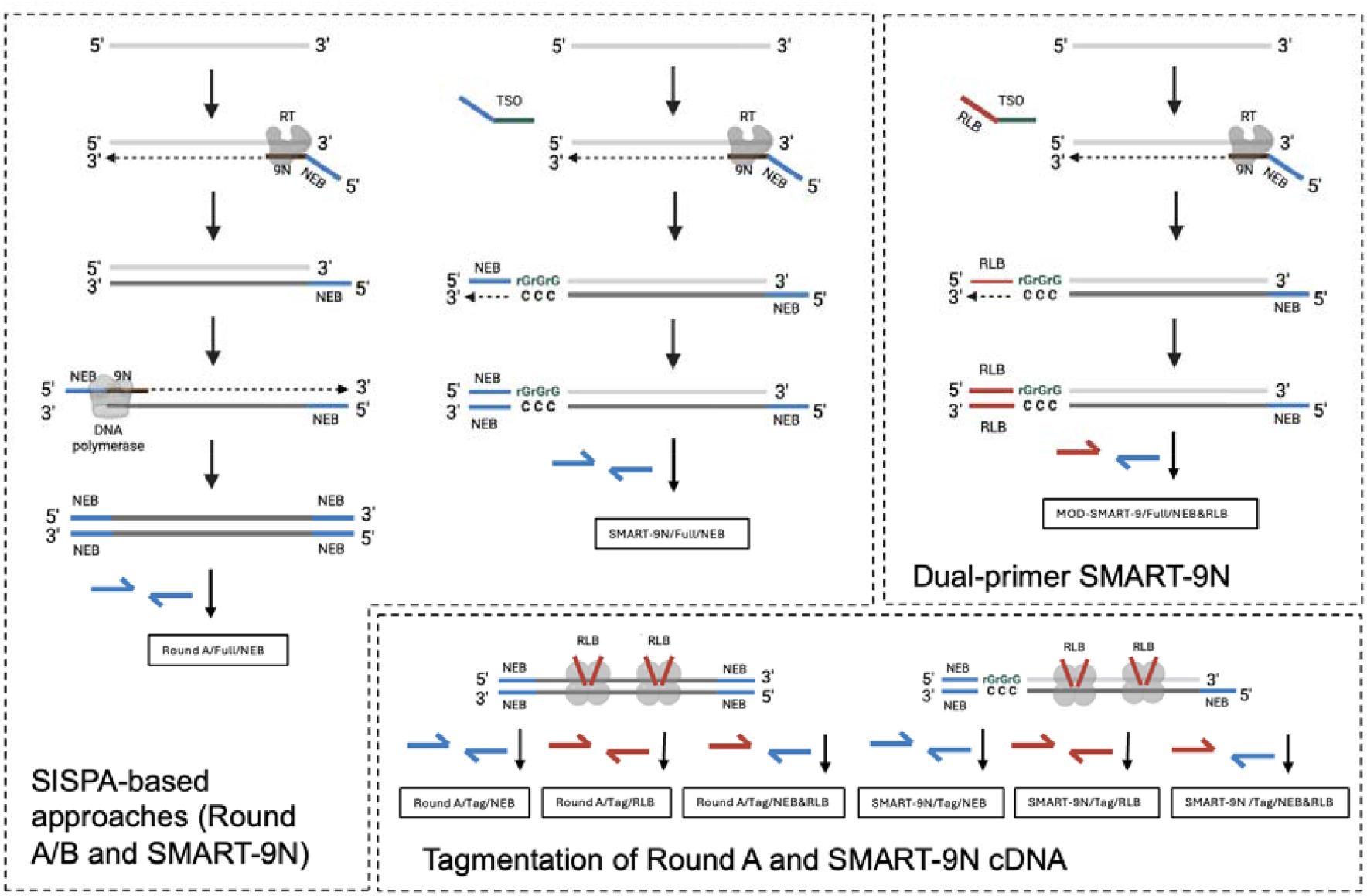
Schematic showing the different approaches used in this study and the names given for each of the conditions tested. ‘SISPA-based approaches (Round A/B and SMART-9N)’ shows cDNA synthesis using Round A or SMART-9N followed by single-primer amplification. ‘Tagmentation of Round A and SMART-9N cDNA’ shows tagmentation of cDNA followed by single primer amplification either targeting the adapters introduced during cDNA synthesis (NEB) or by tagmentation (RLB) or both. ‘Dual-primer SMART-9N’ shows a SMART-9N approach with a modified template-switching oligo (TSO) for dual-primer PCR amplification.

#### Impact of input and background using spiked-clinical samples

To investigate the impact of input and background level on performance we spiked the Zeptometrix Respiratory 2.1 panel into clinical extractions selected to have high, medium or low background. This was motivated by concerns about the efficiency of tagmentation as cDNA input quantities when sequencing the Zeptometrix control panel alone were significantly below the recommended input for these protocols. The recommended input for the FRM reaction in the PCR Barcoding Kit 24 V14 is 1-5 ng of DNA ≥4 kb. We were also interested in the enrichment capacity for SISPA-based methods when challenged with higher background levels. These spiked-samples were sequenced using the same eight conditions outlined above.

#### Effect of Single-vs dual-primer PCR for SMART-9N

We modified SMART-9N to produce 5’ full-length cDNA with RLB adapter at the 3’ and NEB adapter at the 5’ end making it compatible with dual-primer PCR. This is referred to as ‘MOD-SMART-9N/Full’. This allowed us to compare single- and dual-primer PCR for cDNA amplification; single-primer PCR being subject to the PS-effect and enabling length-based enrichment whereas dual-primer PCR is not.

#### Control samples

300 µL of Control 1 and 300 µL of Control 2 from the Zeptometrix Respiratory 2.1 panel (Cat. No. NATRPC2.1-BIO, Zeptometrix, USA) were combined to make a more complex community. The Zeptometrix Respiratory 2.1 panel is a qualitative standard containing 19 viral strains (16 RNA and 3 DNA viruses) and 4 bacterial strains, representing common respiratory pathogens. The sample was centrifuged at 10,000 x g for 5 minutes, and subsequently DNase-treated prior to extraction to remove any extracellular DNA to improve viral sensitivity. 10 µL of HL-SAN Triton-free non-specific endonuclease (Cat. No. 70911-202, ArtcticZymes, Norway) was added to 300 µL of supernatant and incubated on a thermomixer at 37 °C for 10 minutes at 1000 rpm. Total nucleic acid was manually extracted using the MagMAX Viral/Pathogen Nucleic Acid Isolation Kit (Cat. No. A4252, Thermo Fisher Scientific, USA) according to the manufacturer’s instructions and eluted in 25 µL of nuclease-free water.

#### Clinical samples

Clinical samples of high, medium and low concentration following pre-treatment were obtained from the NIHR BRC in Birmingham. Extractions of Zeptometrix Respiratory 2.1 panel were spiked into these samples before cDNA synthesis.

#### 9N primer annealing

10 μl of extracted RNA was combined with 1 μl of 12 µM NEB-9N RT primer (5’-AAGCAGTGGTATCAACGCAGAGTACNNNNNNNNN-3’) and 1 μl of 10 mM dNTP mix (Cat. No. R0193, Thermo Scientific, USA), and incubated for 5 min at 65°C, then rapidly cooled on ice for 1 min.

#### SMART-9N cDNA synthesis

12 µL of the annealed RNA from the previous step was mixed with 4 μl of 5X buffer, 1 μL of 0.1 M DTT, 1 μL of SuperScript IV Reverse Transcriptase (Cat. No. 18091200, Thermo Fisher Scientific, USA), 1 μl of RNaseOUT Recombinant Ribonuclease Inhibitor (Cat. No. 10777019, Thermo Scientific, USA), and 1µL of 12 µM template switching oligo NEB-SSP (5’-GCTAATCATTGCAAGCAGTGGTATCAACGCAGAGTACATrGrGrG-3’). The reaction was incubated at 42°C for 90 min, followed by 70°C for 10 min.

#### Round A cDNA synthesis

The first strand was generated by mixing 12 µL of the annealed RNA with 4 μl of 5X buffer, 1 μL of 0.1 M DTT, 1 μL of SuperScript IV Reverse Transcriptase (Cat. No. 18091200, Thermo Fisher Scientific, USA), 1 μl of RNaseOUT Recombinant Ribonuclease Inhibitor (Cat. No. 10777019, Thermo Scientific, USA), and incubating at 42°C for 10 min, followed by 70°C for 10 min. The second strand was synthesised by adding 2 µL of 5X Sequenase buffer, 0.9 µL of the dilution buffer, 0.6 µL of Sequenase T7 DNA polymerase (Cat. No. 70775Y200UN, ThermoFisher, USA), and 7.7 µL of nuclease-free water to the previous reaction, and incubating at 37 ºC for 8 min.

#### Tagmentation

3 µl of cDNA was combined with 1 µl of the Fragmentation Mix (FRM) from the PCR Barcoding Kit 24 V14 (SQK-RPB114.24, Oxford Nanopore Technologies, UK), and incubated at 30°C for 2 min, followed by 80°C for 2 min and 1 min on ice.

#### PCR Amplification

PCR reactions were performed using either the primer corresponding to the ITR adapter sequence, known as ‘NEB’ (5’-AAGCAGTGGTATCAACGCAGAGT-3’) or the primer corresponding to the transposase, known as ‘RLB’ (5’-TTTTTCGTGCGCCGCTTCA-3’) or a combination of both (Figure 1A). PCR reactions were assembled by mixing 12.5 μl of Q5 High-Fidelity 2X Master Mix, (Cat. No. M0492, New England Biolabs, USA), 1 µl of 10 µM PCR primer, 2.5 µL of cDNA and 9 µl of nuclease-free water. Thermocycling conditions were as follows: initial denaturation at 98°C for 45 sec, 30 cycles of 98°C for 15 sec, 62°C for 15 sec, and 65°C for 5 min, and a final extension at 65°C for 10 min.

#### Post-PCR clean-up

20 µl of each PCR reaction was transferred into a 1.5 mL Eppendorf DNA LoBind tube and an equal volume (1x) of AMPure XP Beads (Cat. No. A63880, Beckman Coulter, USA) was added, incubated for 5 minutes, and washed twice with 200 µl of freshly prepared 70% ethanol. The samples were eluted in 20 µl of nuclease-free water.

#### Library Preparation and Sequencing

Cleaned-up PCR products were barcoded using the ARTIC LoCost Amplicon Sequencing Protocol (SQK-NBD114) (https://www.protocols.io/view/artic-locost-amplicon-sequencing-protocol-sqk-nbd1-5jyl885p7l2w/v1) . Sequencing was performed on a PromethION P2i instrument using R10.4.1 flow cells for 72 hours.

#### Bioinformatic Analysis

Breadth and depth of genome coverage were estimated with k-mer containment analysis using Skope 0.3.0 (https://doi.org/10.5281/zenodo.20185515). Draft reference genomes were assembled, polished, and published (https://doi.org/10.5281/zenodo.16412856) for 16/19 viral strains in the Zeptometrix Respiratory Panel 2.1. Genome containment was estimated using densely sampled reference genome k-mers. Containment values represent the proportion of 31-mers sampled for each reference genome detected in the sequencing data meeting an indicated abundance threshold of 10. The highest yielding replicate for each condition was analysed. Reads of human origin were dehosted with Deacon 0.15.0 (Constantinides et al. 2025). A code notebook enabling reproduction of key results and figures from published raw sequence data is available (https://zenodo.org/records/20185851).

## Results

### Adapter introduction strategy determines RNA virus sensitivity

We evaluated eight conditions representing combinations of two cDNA synthesis methods (SMART-9N and Round A), two adapter introduction strategies (during cDNA synthesis and via tagmentation), and two amplification strategies (single-primer PCR using NEB, RLB primers or both) (**Figure 1**). The four best-performing conditions shared a common feature: use of the NEB primer, which amplifies adapters introduced during cDNA synthesis. SMART-9N/Full/NEB and SMART-9N/Tag/NEB recovered 15/16 and 16/16 viral genomes respectively, while Round A/Full/NEB and Round A/Tag/NEB recovered 13/16 and 11/16 (**Table 1**). None of the conditions relying on the RLB primer, which amplifies adapters introduced by tagmentation, recovered any viral genomes.

**Table 1.** Summary table for the investigation into adapter insertion and amplification strategy. The yields were calculated using `skope classify` for unambiguously classified reads within 1Gbp samples of the highest yielding replicate per condition. Percentages were calculated by dividing the number of bases classified for each taxa by the total yield multiplied by 100. Number of viral strains with 0.5 containment at depth ≥10 out of 16 total. Rows are sorted by descending viral yield percentage.

| Condition | Total yield (Mbp) | Human yield (Mbp) | Human yield (%) | Viral yield (Mbp) | Viral yield (%) | Genomes recovered |
| --- | --- | --- | --- | --- | --- | --- |
| SMART-9N/Tag/NEB | 506 | 192 | 38% | 119 | 24% | 16 |
| SMART-9N/Full/NEB | 605 | 240 | 40% | 139 | 23% | 15 |
| Round A/Full/NEB | 1,000 | 331 | 33% | 225 | 22% | 13 |
| Round A/Tag/NEB | 1,000 | 327 | 33% | 190 | 19% | 11 |
| SMART-9N/Tag/RLB | 4 | 1 | 13% | 0 | 7% | 0 |
| SMART-9N/Tag/NEB&RLB | 15 | 1 | 9% | 1 | 6% | 0 |
| Round A/Tag/NEB&RLB | 18 | 0 | 0% | 0 | 0% | 0 |
| Round A/Tag/RLB | 36 | 0 | 0% | 0 | 0% | 0 |

There was similar performance between tagmented and non-tagmented cDNA when amplified with the NEB primer (e.g. SMART-9N/Tag/NEB versus SMART-9N/Full/NEB. For Round A, the non-tagmented cDNA (Round A/Full/NEB) performed slightly better, recovering two more genomes than the tagmented cDNA (Round A/Tag/NEB). This may indicate that tagmentation of DNA is more efficient than tagmentation of RNA/DNA heteroduplexes such as those produced by SMART-9N. To determine whether transposase activity was inhibited in the cDNA buffer, we included a SPRI clean-up step after cDNA synthesis prior to tagmentation; this did not rescue performance in any RLB-amplified condition (**Supplementary Figure 1**), suggesting that inhibition of transposase activity alone does not account for the failure of RLB-amplified conditions.

Conditions where tagmented cDNA was amplified using RLB (Round A/Tag/RLB, Round A/Tag/NEB&RLB, SMART-9N/Tag/RLB and SMART-9N/Tag/NEB&RLB) produced very low sequencing yields with predominantly unclassified reads. For all other conditions, between 33 and 40% of reads were classified as human, likely deriving from cell lines used for *in vitro* culture of the viral strains in the panel. SMART-9N/Full/NEB generated the highest proportion of viral sequences at 24% (**Figure 2**). There were differences in taxonomic composition between the two SISPA-based approaches: Round A generated a higher proportion of bacterial sequences while SMART-9N produced more DNA viral sequences. SMART-9N longer read lengths than Round A, with a higher mode and longer tail extending to 2 kb. Tagmentation appeared to have no effect on read length for SMART-9N and the read lengths assigned to different classes were similar (**Supplementary Figure 2**).

**Figure 2.**
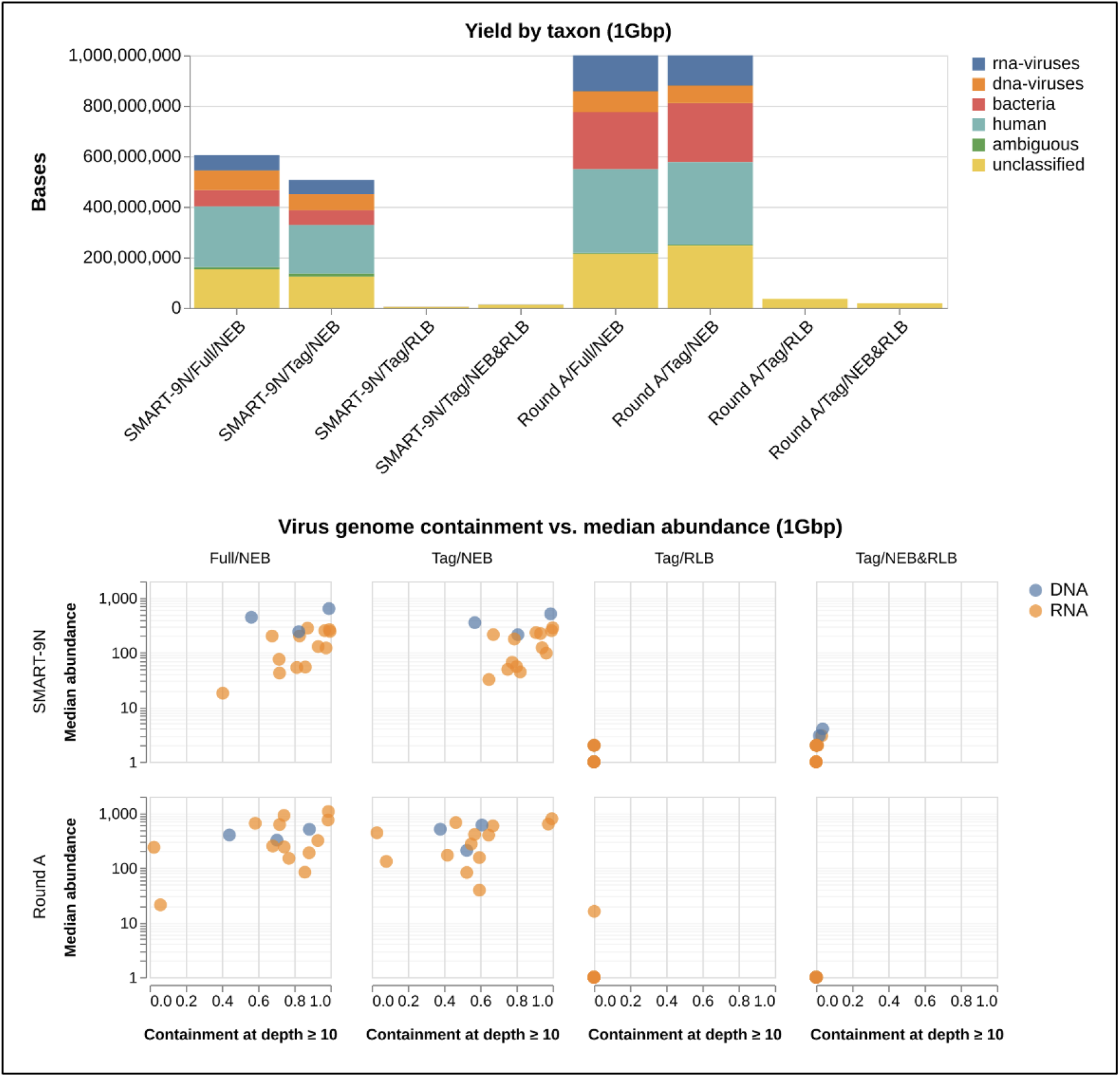
Impact of adapter introduction and amplification strategy on taxonomic composition and viral genome containment and abundance. **Top panel**. Taxonomic composition by kingdom for each condition. Unclassified means no k-mers were identified and ambiguous means k-mers were identified across more than one class. **Bottom panel**. Viral genome containment at depth ≥10 versus median abundance for each viral species in 1 Gbp subsamples. Each point represents one viral species for RNA viruses (orange) and DNA viruses (blue).

**Figure 2.**
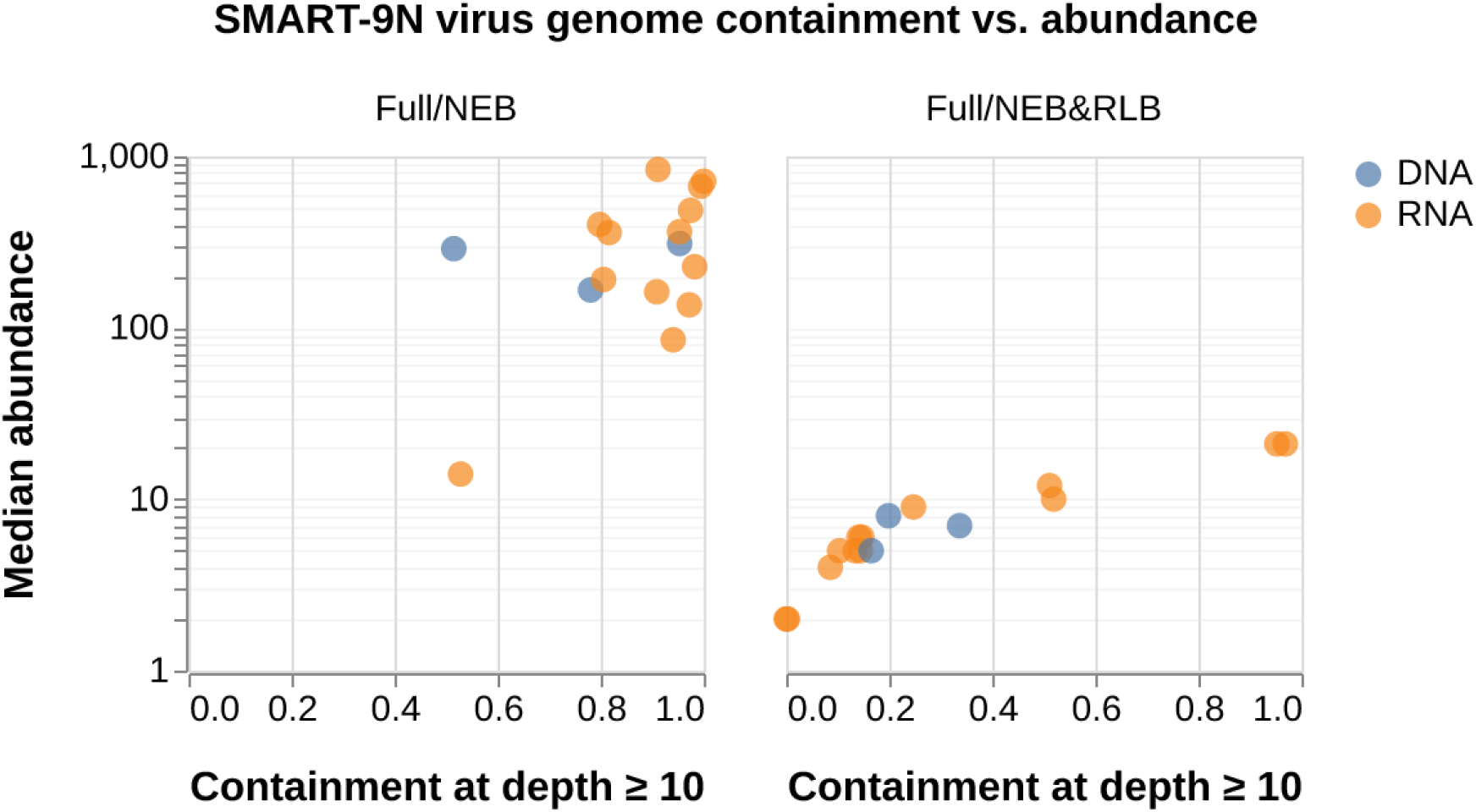
Scatter plots showing the effect of single- and dual-primer amplification on viral genome containment and abundance for different background levels. Viral genome containment at depth ≥10 versus median abundance for each viral species in 1 Gbp subsamples. Each point represents one viral species for RNA viruses (orange) and DNA viruses (blue). Highest yielding replicate only for each condition, does not include controls.

SISPA-based approaches (SMART-9N/Full/NEB and Round A/Full/NEB) produced high viral containment and abundance for all viruses in the panel, with median abundances between 10 and 1000 consistent with a broad range of viral titres in the input sample. Containment was marginally higher for SMART-9N, potentially indicating greater cDNA complexity (**Figure 2**). SARS-CoV-2 and HCoV-229E showed notably lower containment for Round A than SMART-9N, though batch-to-batch variation in the control panel is a known confounding factor.

### SISPA-based approaches outperform tagmentation at different background levels

To investigate performance across a range of clinically relevant host backgrounds, we spiked the Zeptometrix Respiratory 2.1 panel into clinical extractions representing low, medium and high background levels and applied the same eight conditions described above. Background level had a significant impact on viral recovery across all conditions, with best performance in the low background setting and worst in the high background setting **(Table 2)**. SMART-9N/Full/NEB and SMART-9N/Tag/NEB produced 17 and 20% viral yields (lower than when sequencing just the control) yet still managed to recover 15 genomes. At medium background the viral yield was reduced to 1% and the number of genomes recovered was 5. Here there was a noticeable difference between SMART-9N and Round A cDNA which only recovered 7 and 3 genomes for the SMART-9N/Full/NEB and SMART-9N/Tag/NEB conditions at low background and none at medium background. In the high background, no condition recovered any genomes demonstrating that the background was sufficiently high it could not be overcome by length-based enrichment alone.

**Table 2.** Summary table for the investigation into the impact of input and background using spiked clinical samples. The yields were calculated using `skope classify` for unambiguously classified reads within 1Gbp samples of the highest yielding replicate per condition. Percentages were calculated by dividing the number of bases classified for each taxa by the total yield multiplied by 100. Number of viral strains with 0.5 containment at depth ≥10 out of 16 total. Rows are sorted by condition and background.

| Condition | Host background | Total yield (Mbp) | Human yield (Mbp) | Human yield (%) | Viral yield (Mbp) | Viral yield (%) | Genomes recovered |
| --- | --- | --- | --- | --- | --- | --- | --- |
| SMART-9N/Full/NEB | low | 1,000 | 343 | 34% | 174 | 17% | 15 |
| SMART-9N/Full/NEB | medium | 1,000 | 959 | 96% | 6 | 1% | 5 |
| SMART-9N/Full/NEB | high | 1,000 | 992 | 99% | 0 | 0% | 0 |
| SMART-9N/Tag/NEB | low | 1,000 | 330 | 33% | 199 | 20% | 15 |
| SMART-9N/Tag/NEB | medium | 1,000 | 958 | 96% | 6 | 1% | 5 |
| SMART-9N/Tag/NEB | high | 1,000 | 991 | 99% | 0 | 0% | 0 |
| SMART-9N/Tag/RLB | low | 50 | 1 | 2% | 0 | 0% | 0 |
| SMART-9N/Tag/RLB | medium | 40 | 5 | 14% | 0 | 0% | 0 |
| SMART-9N/Tag/RLB | high | 1,000 | 971 | 97% | 0 | 0% | 0 |
| SMART-9N/Tag/NEB&RLB | low | 128 | 2 | 1% | 0 | 0% | 0 |
| SMART-9N/Tag/NEB&RLB | medium | 275 | 77 | 28% | 0 | 0% | 0 |
| SMART-9N/Tag/NEB&RLB | high | 1,000 | 928 | 93% | 0 | 0% | 0 |
| Round A/Full/NEB | low | 1,000 | 355 | 35% | 158 | 16% | 7 |
| Round A/Full/NEB | medium | 1,000 | 988 | 99% | 2 | 0% | 0 |
| Round A/Full/NEB | high | 1,000 | 981 | 98% | 0 | 0% | 0 |
| Round A/Tag/NEB | low | 1,000 | 341 | 34% | 169 | 17% | 3 |
| Round A/Tag/NEB | medium | 1,000 | 983 | 98% | 3 | 0% | 0 |
| Round A/Tag/NEB | high | 1,000 | 985 | 98% | 0 | 0% | 0 |
| Round A/Tag/RLB | low | 8 | 1 | 19% | 0 | 1% | 0 |
| Round A/Tag/RLB | medium | 12 | 2 | 19% | 0 | 0% | 0 |
| Round A/Tag/RLB | high | 1,000 | 995 | 99% | 0 | 0% | 0 |
| Round A/Tag/NEB&RLB | low | 35 | 4 | 12% | 0 | 0% | 0 |
| Round A/Tag/NEB&RLB | medium | 53 | 9 | 17% | 0 | 0% | 0 |
| Round A/Tag/NEB&RLB | high | 1,000 | 949 | 95% | 0 | 0% | 0 |

Assignments to human reads increased with background level across all conditions. For SMART-9N/Full/NEB the yield assigned to human was 34% (low), 96% (medium) and 99% (high). For the same condition the proportion of sequences assigned to viruses was 17% (low), 1% (medium) and 0% (high) background respectively. For Round A the yield assigned to human was 35% (low), 99% (medium) and 98% (high) while the proportion of sequences assigned to viruses was 16% (low), 0% (medium) and 0% (high) background. This indicates that SMART-9N provided a stronger enrichment effect.

In this investigation, tagmented cDNA amplified by RLB produced a high sequencing yield for the high background but not the low or medium. This suggests that input from the Zeptometrics panel alone was inadequate to produce amplifiable inputs, either due to the overall mass or the molecular weight. Despite the higher yield, no condition using tagmentation amplified by RLB recovered any viruses across any background setting. In two instances, high background samples produced unexpectedly elevated abundance (∼100x) for individual viruses at low containment, which may represent efficient amplification of a low complexity input and warrants further investigation **(Supplementary Figure 3)**.

### Single-primer PCR is essential for suppression-based enrichment

To directly test the contribution of single-primer PCR to PS-effect-based enrichment, independent of adapter introduction strategy, we compared single- and dual-primer amplification of full-length SMART-9N cDNA using a modified protocol (MOD-SMART-9N/Full) in which the template-switching oligo was modified to incorporate the RLB adapter at the 3’ end and the NEB adapter at the 5’ end, making the cDNA compatible with dual-primer PCR. Single-primer PCR produced high abundance and containment for all viral genomes, whereas dual-primer PCR produced low abundance for all genomes (**Figure 2**). Two viral genomes (HPIV2 and HPIV3) still had high containment even at low abundance potentially indicating a high complexity cDNA library yet lack of PCR-based enrichment. This confirms that single-primer amplification, and the PS-effect it enables, is the critical determinant of viral enrichment even when tagmentation efficiency is input dependent.

## Discussion

### The PCR suppression effect provides a mechanistic basis for SISPA-based viral enrichment

Our results demonstrate that library preparation strategy is a critical determinant of RNA virus sensitivity in clinical metagenomics. The complete failure of RLB-amplified conditions to recover any viral genomes, regardless of cDNA synthesis method, is consistent with our mechanistic model: tagmentation fragments cDNA to a uniform size distribution, eliminating the length differential between host and viral molecules that enables PS-effect-based enrichment. Conversely, SISPA-based methods are highly effective at sequencing viruses even when titres are low, with titres reported to range between Ct 26.6 and 39.1 in the Zeptometrix Respiratory 2.1 panel (Doyle et al. 2025).

The similar performance of SMART-9N/Tag/NEB and SMART-9N/Full/NEB is likely due to the fact that transposition is inefficient for RNA/DNA heteroduplex substrates meaning there is little effect on the amplification. The slight reduction in containment seen for Round A following tagmentation, but not for SMART-9N, may indicate that Round A cDNA is tagmented more efficiently than SMART-9N but that there are still sufficient untagmented molecules available for amplification. The failure of the SPRI clean-up to rescue RLB performance argues against simple transposase inhibition in the cDNA buffer as the primary explanation for RLB failure. This supports the interpretation that the mechanism is not one of failed tagmentation but of successful tagmentation eliminating the size distribution required for enrichment.

There were notable differences in taxonomic composition between SMART-9N and Round A. Round A generated a higher proportion of bacterial sequences while SMART-9N produced more DNA viral sequences, and SMART-9N produced marginally longer reads. These differences likely reflect the different molecular biology used - template switching in SMART-9N versus Sequenase-based second strand synthesis in Round A - which results in differences in composition and read-length. While both approaches are effective for RNA virus detection, these differences may be relevant when designing protocol for clinical metagenomics.

### Host background defines the upper limit of suppression-based viral enrichment

The spiked clinical sample investigation demonstrated how host background level is one of the main limitations to sensitivity in clinical metagenomics, something which has been widely reported (Benoit et al. 2024). At low background, SMART-9N/Full/NEB and SMART-9N/Tag/NEB maintained reasonable viral yields and recovered 15 genomes despite the additional host complexity relative to the control-only experiment. However, at medium background viral yield collapsed to 1% and genome recovery fell to 5, and at high background no condition recovered any viral genomes, demonstrating that beyond a critical threshold of host background, length-based enrichment alone is insufficient to rescue viral signal.

A performance differential between SMART-9N and Round A was seen at low and medium background which was not observed in the control-only experiment. SMART-9N/Full/NEB and SMART-9N/Tag/NEB recovered 7 and 3 genomes respectively at low background where Round A recovered none, and this advantage was maintained at medium background. This suggests that SMART-9N may have a more powerful enrichment effect possibly due to generating longer cDNA and therefore a bigger size differential or that it generates a more complex cDNA library than Round A due to the smaller volume of the template-switching reaction. The practical implication is that SMART-9N may be the preferred approach for clinical samples where low viral titre and moderate host background are anticipated simultaneously.

The behaviour of tagmented cDNA amplified by RLB differed markedly between the control-only and spiked clinical experiments. In the control-only experiment RLB-amplified conditions produced very low yields, while in the high background spiked samples it produced substantially higher yields. This is consistent with the input mass of the low and medium backgrounds being too low for efficient tagmentation. Despite this improved yield, no RLB-amplified condition recovered any viral genomes across any background setting, reinforcing that the failure of tagmentation-based approaches is a consequence of the loss of size-based enrichment rather than simply insufficient input for the tagmentation reaction.

The anomalous finding of high abundance at low containment for individual viruses in high background RLB-amplified samples could reflect efficient amplification of a low-complexity input, where a small number of molecules are heavily duplicated during PCR, is consistent with stochastic behaviour at the limits of sensitivity (**Supplementary Figure 3**). This highlights the importance of using containment rather than abundance alone as the primary metric for assay performance, as abundance in isolation can be misleading in low-complexity libraries.

### Single-primer PCR is essential for size-based enrichment

The direct comparison of single- and dual-primer PCR using modifications to the SMART-9N protocol is strong evidence for the PS-effect based enrichment model proposed here. By controlling other variables, we demonstrate that single-primer PCR is critical for viral enrichment and provides strong evidence that, regardless of tagmentation efficiency due to input or inhibition, the PS-effect is driving viral enrichment. Without single-primer PCR the libraries likely reflect the underlying abundance of the sample rather than the viral abundances achievable with PCR suppression-based enrichment, dramatically reducing sensitivity. This has practical implications beyond the protocols tested here. Any modification that introduces a second primer into the amplification step, whether through a modified template-switching oligo, a dual-adapter tagmentation kit, or a barcoding strategy requiring two primers, will abrogate the enrichment benefit of SISPA-based amplification. This principle extends to short-read sequencing approaches: RNA fragmentation by chemical means, as used in Illumina TruSeq Stranded Total RNA and KAPA RNA HyperPrep, produces a uniform size distribution analogous to tagmentation and will similarly prevent PS-effect-based enrichment. SMARTer Stranded Total RNA-Seq preserves size distribution but requires two-primer PCR to maintain strand information, meaning it will also not benefit from suppression-based enrichment. These considerations suggest that the performance advantages of SISPA-based approaches are not limited to long-read sequencing and warrant evaluation in short-read clinical metagenomics workflows.

### Implications for widely used clinical metagenomics protocols

Despite the limitations identified here, tagmentation of cDNA is employed in a number of widely used clinical metagenomics protocols, including the UCSF CSF mNGS assay (Wilson et al. 2019; Miller et al. 2019) and the Pan-microbial protocol (Alcolea-Medina et al. 2024). The apparent success of these protocols in clinical validation studies likely reflects two factors. First, many validation studies have been conducted using sample types with relatively low host background, where the advantage of PS-effect-based enrichment is less critical and high sequencing depth can compensate for reduced enrichment efficiency. Second, validation using clinical specimens with unknown or variable viral titres makes it difficult to detect sensitivity differences that would be apparent under controlled conditions. The data from the multicentre benchmarking study by Lopez-Labrador et al. (2024), in which several protocols included tagmentation of cDNA (Grädel et al. 2022; Atkinson et al. 2023; Kufner et al. 2019) showed lower normalised read counts than SISPA-based methods (Wollants et al. 2020; Vanmechelen et al. 2022), is consistent with our findings. It is important to note that SISPA-based preamplification followed by tagmentation (Greninger et al. 2015; Xu et al. 2020; Xu et al. 2021; Lewandowska et al. 2015; Fernandez-Cassi et al. 2018; Kramná & Cinek 2018; Gauthier et al. 2024; Sanderson et al. 2026) or ligation (Wollants et al. 2020; Vanmechelen et al. 2022) or combined amplification and library preparation (Pichler et al. 2023; Claro et al. 2021) are distinct and valid approaches. In this case, size-based enrichment has already occurred during the SISPA amplification step. Our findings argue against direct tagmentation of cDNA, not against tagmentation as a library preparation entirely.

### Limitations

The PCR suppression-based enrichment mechanism has important limitations. Viruses with small genomes, such as parvoviruses and certain picornaviruses, may experience stronger suppression than larger viruses, potentially reducing sensitivity for these targets. Multi-segmented viruses such as influenza have individual segments short enough to limit enrichment potential; the NS and M segments of Influenza A are particularly relevant in this regard. These limitations should be considered when interpreting negative results for small or segmented RNA viruses in SISPA-based protocols. While the Zeptometrix Respiratory 2.1 panel provides a convenient and reproducible reference standard, it does not fully represent the complexity of clinical specimens. The viral strains are produced by *in vitro* cell culture, inactivated, and resuspended in a proprietary buffer, and different batches are not guaranteed to have the same viral titres. Validation in clinically realistic matrices with quantified viral titres spiked into human cell backgrounds remains an important next step. Single-primer PCR produces higher rates of chimeric reads than dual-primer PCR. These chimeras require appropriate bioinformatic handling and may complicate variant analysis or metagenomic assembly. Containment-based analysis approaches, as used here, are better suited to handling chimeric reads than alignment or assembly-based approaches, and we recommend their use in conjunction with SISPA-based protocols. DNA viruses were present in the panel but were not the primary focus of this analysis. DNA viruses may be detected through amplification of RNA intermediates during active replication and we cannot distinguish these with the experiments presented here.

### Practical recommendations

Based on our findings, we recommend the following for clinical metagenomic protocols targeting RNA viruses:

- **Use SISPA-based methods where RNA virus sensitivity is important:** Both SMART-9N and Round A approaches have high sensitivity for RNA viruses. SMART-9N offers a simpler single-tube reaction, while Round A/B is quicker.
- **Apply tagmentation only to SISPA-amplified products:** Tagmentation-based library preparation should be used on SISPA amplified products, which have already benefited from size-based enrichment. They should not be used directly on cDNA inputs.
- **Combine with DNase pretreatment:** Pre-extraction DNase treatment degrades extracellular host DNA, improving viral sensitivity, whilst preserving DNA virus genomes for sequencing via the SISPA-Seq mechanism.
- **Dual-arm protocols for comprehensive detection:** For pathogen agnostic clinical metagenomics, parallel processing with viral or bacterial specific protocol arms will likely outperform combined protocols.
- **Implement chimera-aware bioinformatic pipelines:** Single-primer PCR increases the rate of chimera formation, and bioinformatic methods should accommodate this. Lowest common ancestor (LCA) read classification approaches such as Kraken may report artificially high proportions of unclassified reads due to chimeras.
- **Validate protocols using appropriate controls:** Mock communities containing RNA and DNA virus at known titres, spiked into human cell matrices should be used to validate pathogen agnostic protocols. It should not be assumed that protocols will work equally well for different pathogens.

### Future perspectives

These results strongly suggest that SISPA-based approaches have the most potential for further development, with improvements likely to extend detection to lower viral loads, earlier infection time points, and sample types currently considered challenging. The relationship between viral genome size, segmentation, and PS-effect-based enrichment warrants quantitative characterisation, a systematic analysis across a broader range of viral genome sizes would allow performance expectations to be defined for different viral families and identify the lower size threshold below which suppression becomes counterproductive rather than beneficial. SISPA could be optimised either to amplify small or segmented RNA viruses or by increasing enrichment by increasing the length differential. Given the importance to public health, optimising for sensitivity, even at the cost of increased workflow complexity, represents the priority for surveillance applications.

## Data Availability

Resources and instructions for downloading raw reads and replicating results in this manuscript are available from https://github.com/bede/2026-protocol-optimisation.

https://github.com/bede/2026-protocol-optimisation

## Data availability

### Ethics

The study from which the clinical samples were obtained received ethical approval from the Northwest – Haydock Research Ethics Committee (Ref: 23/NW/0103) and the Health Research Authority (Ref: 323039).

## References

Alcolea-Medina, A. et al., 2024. Unified metagenomic method for rapid detection of microorganisms in clinical samples. Communications Medicine, 4(1), p.135.

Allander, T. et al., 2001. A virus discovery method incorporating DNase treatment and its application to the identification of two bovine parvovirus species. Proceedings of the National Academy of Sciences of the United States of America, 98(20), pp.11609–11614.

Atkinson, L. et al., 2023. Untargeted metagenomics protocol for the diagnosis of infection from CSF and tissue from sterile sites. Heliyon, 9(9), p.e19854.

Benoit, P. et al., 2024. Seven-year performance of a clinical metagenomic next-generation sequencing test for diagnosis of central nervous system infections. Nature Medicine, 30(12), pp.3522–3533.

Bohlander, S.K. et al., 1992. A method for the rapid sequence-independent amplification of microdissected chromosomal material. Genomics, 13(4), pp.1322–1324.

Charalampous, T. et al., 2019. Nanopore metagenomics enables rapid clinical diagnosis of bacterial lower respiratory infection. Nature Biotechnology, 37(7), pp.783–792.

Claro, I.M. et al., 2021. Rapid viral metagenomics using SMART-9N amplification and nanopore sequencing. Wellcome Open Research, 6, p.241.

Constantinides, B., Lees, J. & Crook, D., 2025. Deacon: fast sequence filtering and contaminant depletion. bioRxiv. Available at: 10.1101/2025.06.09.658732.

Dai, Z.-M. et al., 2007. PCR-suppression effect: kinetic analysis and application to representative or long-molecule biased PCR-based amplification of complex samples. Journal of Biotechnology, 128(3), pp.435–443.

Doyle, R. et al., 2025. Optimisation of whole cell human depletion provides increased sensitivity and microbial genome coverage from respiratory metagenomic assays. medRxiv. Available at: 10.1101/2025.10.10.25337716.

Feehery, G.R. et al., 2013. A method for selectively enriching microbial DNA from contaminating vertebrate host DNA. PloS One, 8(10), p.e76096.

Fernandez-Cassi, X., Rusiñol, M. & Martínez-Puchol, S., 2018. Viral concentration and amplification from human serum samples prior to application of next-generation sequencing analysis. Methods in Molecular Biology (Clifton, N.J.), 1838, pp.173–188.

Froussard, P., 1992. A random-PCR method (rPCR) to construct whole cDNA library from low amounts of RNA. Nucleic Acids Research, 20(11), p.2900.

Gauthier, N.P.G. et al., 2024. Validation of an automated, end-to-end metagenomic sequencing assay for agnostic detection of respiratory viruses. The Journal of Infectious Diseases, 230(6), pp.e1245–e1253.

Grädel, C. et al., 2022. Genome sequences of rare human Enterovirus genotypes recovered from clinical respiratory samples in Bern, Switzerland. Microbiology Resource Announcements, 11(9), p.e0027622.

Greninger, A.L. et al., 2015. Rapid metagenomic identification of viral pathogens in clinical samples by real-time nanopore sequencing analysis. Genome Medicine, 7(1), p.99.

Gu, W. et al., 2016. Depletion of Abundant Sequences by Hybridization (DASH): using Cas9 to remove unwanted high-abundance species in sequencing libraries and molecular counting applications. Genome Biology, 17(1), p.41.

Hasan, M.R. et al., 2016. Depletion of human DNA in spiked clinical specimens for improvement of sensitivity of pathogen detection by next-generation sequencing. Journal of Clinical Microbiology, 54(4), pp.919–927.

Karpinets, T.V. et al., 2006. RNA:protein ratio of the unicellular organism as a characteristic of phosphorous and nitrogen stoichiometry and of the cellular requirement of ribosomes for protein synthesis. BMC Biology, 4(1), p.30.

Kim, M. et al., 2024. Host DNA depletion on frozen human respiratory samples enables successful metagenomic sequencing for microbiome studies. Communications Biology, 7(1), p.1590.

Kramná, L. & Cinek, O., 2018. Virome sequencing of stool samples. Methods in Molecular Biology (Clifton, N.J.), 1838, pp.59–83.

Kufner, V. et al., 2019. Two years of viral metagenomics in a tertiary diagnostics unit: Evaluation of the first 105 cases. Genes, 10(9), p.661.

Lewandowska, D.W. et al., 2015. Unbiased metagenomic sequencing complements specific routine diagnostic methods and increases chances to detect rare viral strains. Diagnostic Microbiology and Infectious Disease, 83(2), pp.133–138.

Liu, B. et al., 2020. An optimized metagenomic approach for virome detection of clinical pharyngeal samples with respiratory infection. Frontiers in Microbiology, 11, p.1552.

Longhi, G. et al., 2024. Saponin treatment for eukaryotic DNA depletion alters the microbial DNA profiles by reducing the abundance of Gram-negative bacteria in metagenomics analyses. Microbiome Research Reports, 3(1), p.4.

Marotz, C.A. et al., 2018. Improving saliva shotgun metagenomics by chemical host DNA depletion. Microbiome, 6(1), p.42.

Miller, S. et al., 2019. Laboratory validation of a clinical metagenomic sequencing assay for pathogen detection in cerebrospinal fluid. Genome Research, 29(5), pp.831–842.

Pichler, I. et al., 2023. Rapid and sensitive single-sample viral metagenomics using Nanopore Flongle sequencing. Journal of Virological Methods, 320(114784), p.114784.

Reyes, G.R. & Kim, J.P., 1991. Sequence-independent, single-primer amplification (SISPA) of complex DNA populations. Molecular and Cellular Probes, 5(6), pp.473–481.

Sanderson, N.D. et al., 2026. Clinical validation of a novel metagenomic nanopore sequencing method for detecting viral respiratory pathogens. medRxiv. Available at: 10.64898/2026.02.06.26345651.

Shagin, D.A. et al., 1999. Regulation of average length of complex PCR product. Nucleic Acids Research, 27(18), p.e23.

Siebert, P.D. et al., 1995. An improved PCR method for walking in uncloned genomic DNA. Nucleic acids research, 23(6), pp.1087–1088.

Vanmechelen, B. et al., 2022. The characterization of multiple novel paramyxoviruses highlights the diverse nature of the subfamily Orthoparamyxovirinae. Virus Evolution, 8(2), p.veac061.

Wang, D. et al., 2003. Viral discovery and sequence recovery using DNA microarrays. PLoS Biology, 1(2), p.E2.

Wilson, M.R. et al., 2019. Clinical metagenomic sequencing for diagnosis of meningitis and encephalitis. The New England Journal of Medicine, 380(24), pp.2327–2340.

Wollants, E. et al., 2020. First genomic characterization of a Belgian Enterovirus C104 using sequence-independent Nanopore sequencing. Infection, Genetics and Evolution: Journal of Molecular Epidemiology and Evolutionary Genetics in Infectious Diseases, 81(104267), p.104267.

Wright, M.S. et al., 2015. SISPA-Seq for rapid whole genome surveys of bacterial isolates. Infection, Genetics and Evolution: Journal of Molecular Epidemiology and Evolutionary Genetics in Infectious Diseases, 32, pp.191–198.

Xu, Y. et al., 2021. Nanopore metagenomic sequencing of influenza virus directly from respiratory samples: diagnosis, drug resistance and nosocomial transmission, United Kingdom, 2018/19 influenza season. Euro surveillance : bulletin Europeen sur les maladies transmissibles [Euro Surveillance : European Communicable Disease Bulletin], 26(27). Available at: http://dx.doi.org/10.2807/1560-7917.ES.2021.26.27.2000004.

Xu, Y. et al., 2020. Nanopore metagenomic sequencing to investigate nosocomial transmission of human metapneumovirus from a unique genetic group among haematology patients in the United Kingdom. The Journal of Infection, 80(5), pp.571–577.

Yu, J. et al., 2024. Comparison of metagenomic next-generation sequencing and blood culture for diagnosis of bloodstream infections. Frontiers in Cellular and Infection Microbiology, 14, p.1338861.

Zhu, Y.Y. et al., 2001. Reverse transcriptase template switching: a SMART approach for full-length cDNA library construction. BioTechniques, 30(4), pp.892–897.

