## Supplementary Data for "Library preparation strategy critically impacts RNA virus sensitivity in clinical metagenomics"

| **Background** | **Sample ID** | **Sample Type** | **Post-extraction concentration** |
| --- | --- | --- | --- |
| Low | BRC 130 | BAL | <0.02 ng/uL |
| Medium | BRC 205 | BAL | 0.1 ng/uL |
| High | BRC156 | ETA | 10.6 ng/uL |

**Supplementary Table 1.** Sample data for extractions used in the investigation into input and background using spiked-clinical samples. Abbreviations used for sample types: BAL (bronchoalveolar lavage), ETA = endotracheal aspirate. Post-extraction concentration as measured by dsDNA HS assay on Qubit 4 fluorimeter.

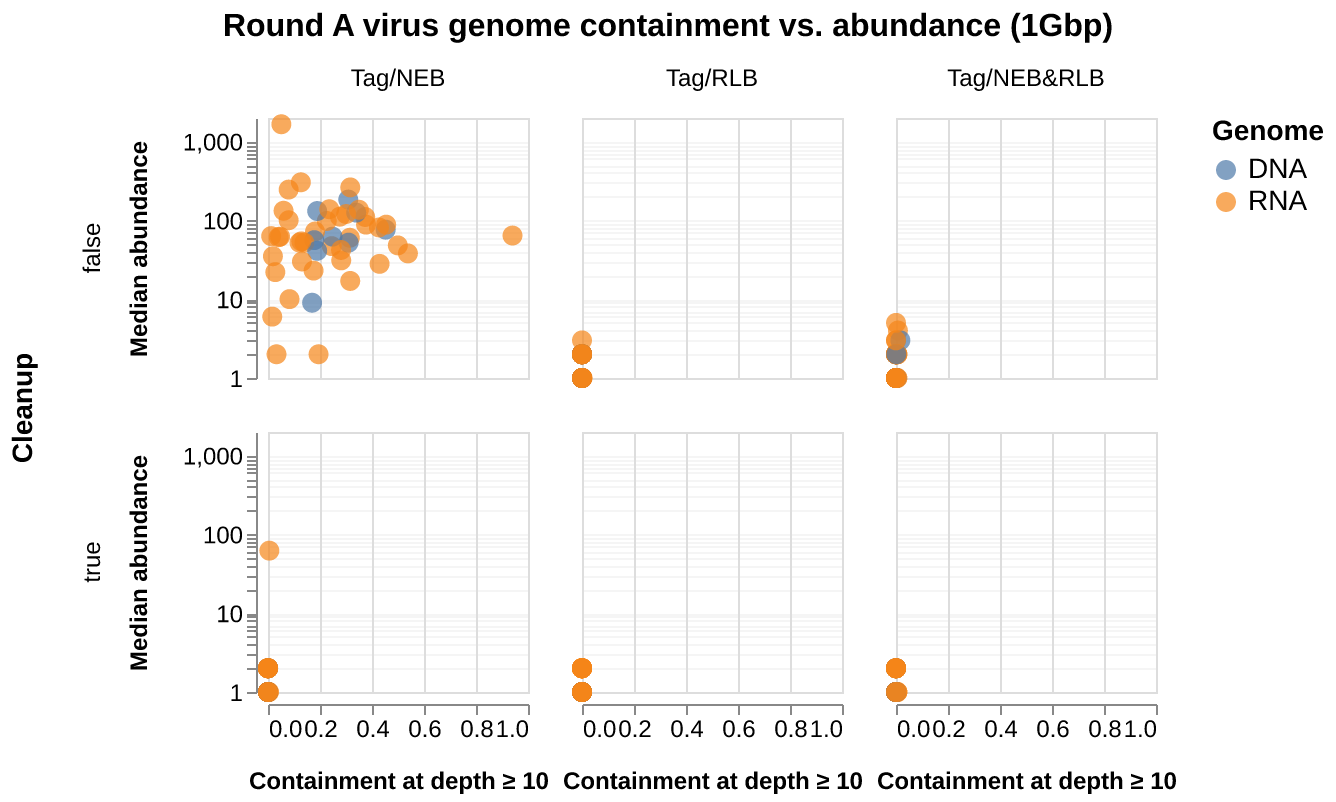

**Supplementary Figure 1.** Scatter plots showing the effect of adapter introduction and amplification strategy on viral genome containment and abundance with and without cDNA clean-up. Viral genome containment at depth ≥10 versus median abundance for each viral species in 1 Gbp subsamples. Each point represents one viral species for RNA viruses (orange) and DNA viruses (blue).

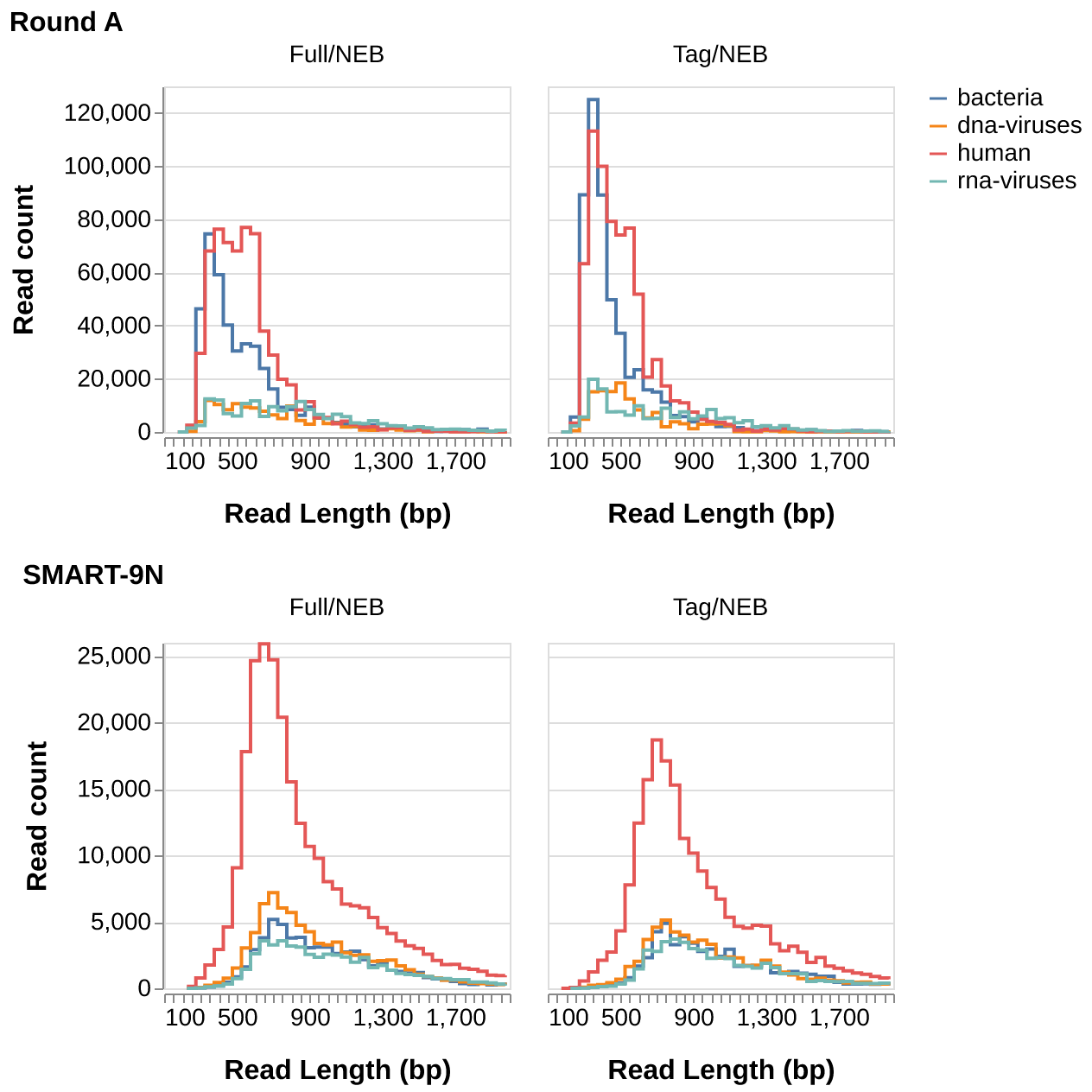

**Supplementary Figure 2.** Read length histograms showing effect of adapter introduction and amplification strategy on read length distributions by kingdom. The read lengths were calculated using skope lenhist for unambiguously classified reads within 1Gbp samples of the highest yielding replicate per condition.

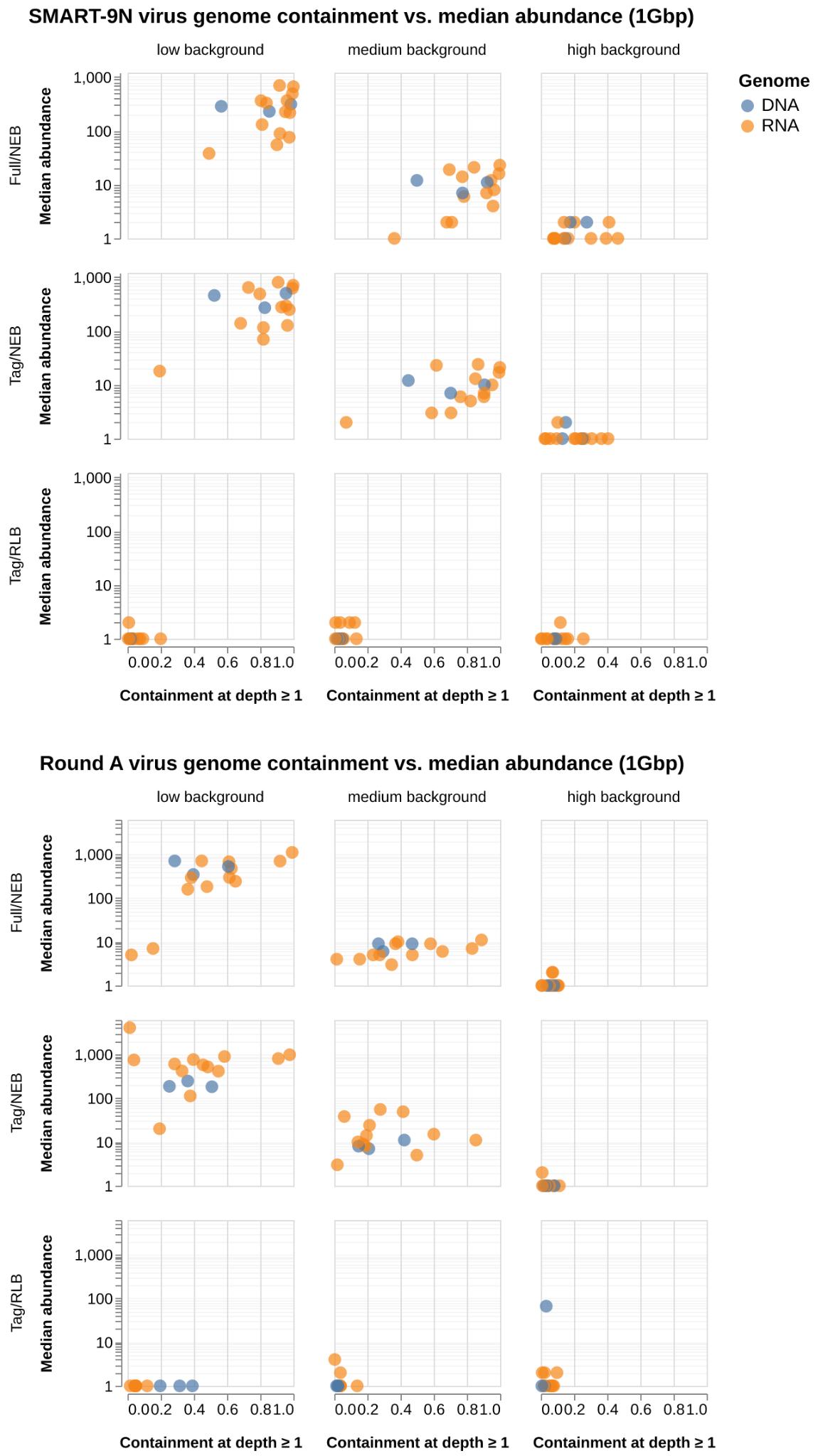

**Supplementary Figure 3.** Scatter plots showing the effect of adapter introduction and amplification strategy on viral genome containment and abundance for different background levels. Viral genome containment at depth ≥1 versus median abundance for each viral species in 1 Gbp subsamples. Each point represents one viral species for RNA viruses (orange) and DNA viruses (blue). Containment at depth ≥1 was used as viral yields were low for medium and high background.
